# RAMDS: Retrieval Augmented Medical Diagnosis System for Explainable Breast Cancer Classification from Ultrasound Images

**DOI:** 10.1101/2024.01.30.24301967

**Authors:** Ethan Thomas Johnson, Jathin K Bande, Johnson Thomas

## Abstract

Breast cancer, a leading cause of cancer-related deaths in women, presents a growing challenge in medical diagnostics. Despite the effectiveness of mammography and ultrasound, the ambiguity in non-invasive scans often necessitates invasive procedures. Our primary goal was to create an AI model that could predict breast cancer with high negative predictive value and reduce unnecessary procedures. This study introduces the Retrieval-Augmented Medical Diagnosis System (RAMDS) for breast cancer, a novel approach combining an AI model with a retrieval-augmented mechanism to enhance diagnostic accuracy and explainability. The RAMDS employs a pretrained ResNet 34 model, fine-tuned on breast ultrasound image datasets from four countries. It integrates a similarity-based weighted adjustment mechanism to compare new cases with historical diagnoses. It’s like having an experienced doctor who remembers every case they’ve ever seen and uses that knowledge to make better decisions. RAMDS improved sensitivity by 11%, and negative predictive value by 9% when compared to the base model. Notably, the RAMDS improves explainability by linking AI predictions to similar historical cases, aligning with the medical community’s interest in transparent and understandable AI decisions.

A unique feature of this system is its adaptability to varied imaging contexts without retraining, addressing the challenge of dataset variability across medical institutions. In conclusion, the RAMDS offers a significant advancement in breast cancer diagnosis, combining enhanced accuracy, explainability, and adaptability. It holds promise for clinical application, though further research is needed to optimize its performance and integrate multi-modal data.

## Introduction

Breast cancer, a leading cause of cancer-related deaths in women, presents a growing challenge in medical diagnostics. Arnold et al. (2022) predicts by 2040 there will be 3 million new cases and 1 million more deaths tied to breast cancer. If detected early, breast cancer has a high cure rate. Hence finding these cancers earlier is of paramount importance. Different imaging modalities like mammography, ultrasound, and MRI can be used for breast cancer screening. Because of high prevalence and mortality, the current guidelines from the U.S. Preventive Services Task Force recommend screening mammograms every 2 years for women between 50 and 74 years of age (Siu, 2016). Mammograms may not be a good option for patients with dense breasts who happen to have a higher risk of developing breast cancer (Boyd, 2007). Breast ultrasounds are preferred in young patients and patients with dense breasts. Ultrasounds are also used when a mass can be felt but cannot be seen on the mammogram. Moreover, breast ultrasounds do not expose patients to radiation, have lower costs, are easily accessible, and can view images in real-time. When compared to mammograms, invasive cancers detected in ultrasounds were node-negative.

As with many medical imaging modalities, there is intra and inter-observer variability in reading breast ultrasounds due to the inherent subjectivity. Reading breast ultrasounds is also time-consuming and prone to errors (Bhowmik, 2022). In one study ultrasounds were found to have low positive predictive value of 4.3% (Berg, 2015). Artificial intelligence (AI) models could reduce the subjectivity, errors, and reading time. There are multiple studies demonstrating radiologist-level performance using AI models for breast ultrasound (Qian, 2021; Kim, 2021). However, most studies were done on limited datasets and from single institutions. A model developed on a local dataset may not perform well on a different dataset collected from another institution or a different ultrasound machine. Most of these models are black boxes, making the decision-making process opaque (Xu et al., 2023).

To address these issues, in this research we aimed to create an explainable AI model with better positive and negative predictive value that could be adapted to local datasets without retraining the whole model.

## Materials and Methods

Our study’s goal was to test whether the novel RAMDS(Retrieval Augmented Medical Diagnosis System), with its unique combination of machine learning and a retrieval-based approach, can outperform a standard AI model in critical aspects of medical diagnostics. The focus is not just on basic accuracy, but also on sensitivity (correctly identifying cancer when it is present), and explainability (providing clear, understandable reasoning behind its diagnoses). By comparing the RAMDS to the base ResNet 34 model, the study seeks to quantify the added value of the retrieval-augmented mechanism in improving breast cancer diagnostics through AI.

### Datasets

All data used in the paper is available in the public domain. Training data consisted of images from BUSC (Rodrigues et al., 2019), BUSI_corrected (Tasnim et al., 2023) and QAMEBI (Ardakani et al., 2023; Hamyoon et al., 2022) datasets. RODTOOK dataset (Kirimasthong et al., 2018) was used for testing. These datasets had breast ultrasound images with corresponding diagnoses. Features of each dataset are listed in Table 1.

**Table 1.**
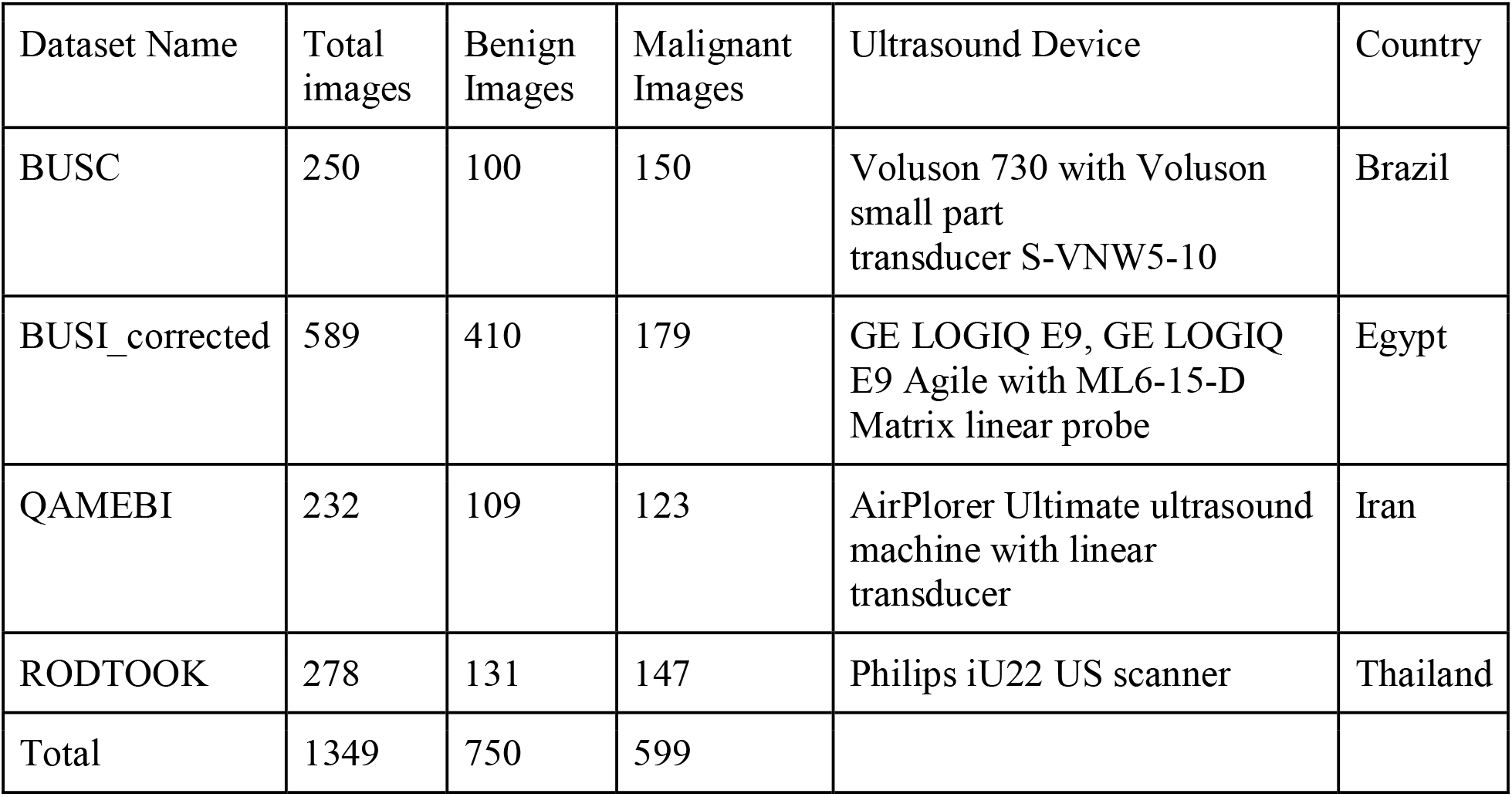
Datasets and features.

Data preprocessing: All images were resized to 256 x 256 pixels and converted to greyscale with padding to maintain aspect ratio. Multiple image augmentation techniques were used to generate more images on the fly for training.

### Model creation

A pretrained ResNet 34 model was fine-tuned on the training dataset using FastAI library. A learning rate finder was used to find the optimal learning rate. Then the learn.fine_tune method was used to fine-tune the model. This ResNet model was used to predict on the test set. After this, the model was used to generate embeddings for the training images, and this was stored in a file. Similar images to the test images were found using cosine similarity between input image embedding and the saved embeddings. N number of similar images and corresponding diagnosis were retrieved.

Then, we created a model that combines a deep learning model’s predictive capability with similarity-based weighted adjustment mechanism, refining the diagnostic threshold based on the concordance with top similar historical cases.

The core of our method lies in the adjusted_prediction function, which operates in several steps:

Base Prediction and Probability: The function begins by obtaining a base prediction and its associated probability from the ResNet34 model.

Similarity Assessment: It then retrieves diagnoses and similarity scores of the top N images closely resembling the input case. These similarities are quantified using cosine similarity measures in the embedding space generated by the same ResNet34 model.

Weighted Agreement Calculation: The function calculates a weighted agreement rate between the base prediction and the retrieved similar cases. More similar cases influence the agreement rate more significantly, assuming that, cases with higher resemblance will provide more relevant contextual information.

Threshold Adjustment: The base threshold, initially set at a specific value, is dynamically adjusted based on the weighted agreement rate. An adjustment factor, determined through a grid search algorithm on the validation test, modulates how much the threshold should shift. The rationale behind this adjustment is to increase the prediction’s confidence when there’s a high agreement with similar cases and decrease it when there’s a discrepancy.

Final Prediction: The adjusted threshold is then applied to the base prediction probability to yield the final diagnosis.

We used Python programming language and packages to conduct statistical analysis. Accuracy, sensitivity, specificity, positive predictive value, negative predictive value, and AUC ROC were calculated to quantify and compare the results.

## Results

The initial training database consisted of 1,071 training images. After removing images with markings inside and images with multiple nodules, there were 900 images in the training set, which included 476 benign images and 424 malignant images. The testing set contained 274 images of which 134 were benign and 140 were malignant.

### ResNet 34 model results

Sensitivity, specificity, accuracy, positive predictive value, and negative predictive value for the ResNet34 model were 79%, 59%, 69%, 67%, and 73% respectively. The AUC for the ResNet34 model was 0.76 (Figure 2).

**Figure 1.**
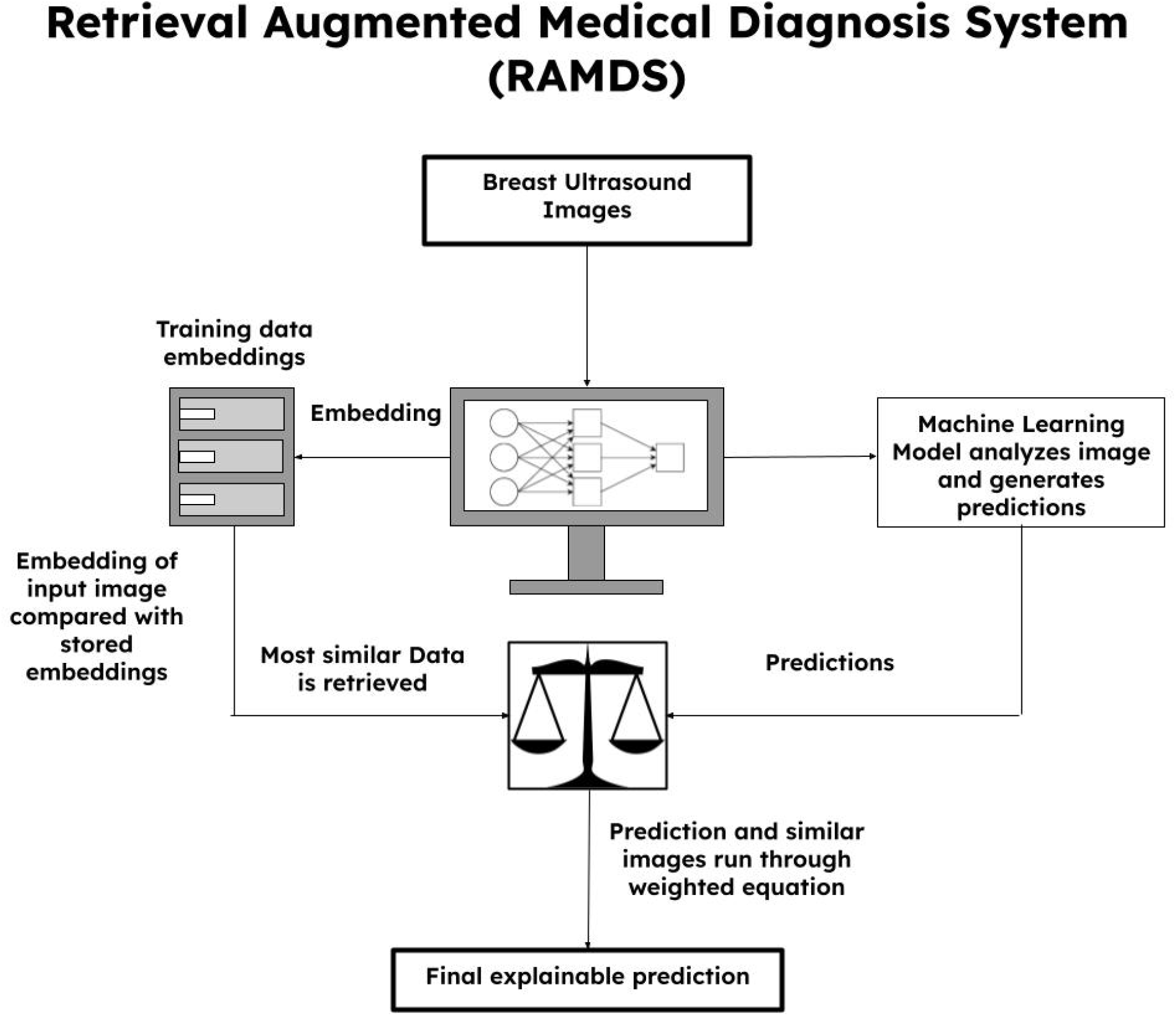
Diagrammatic representation of Retrieval Augmented Medical Diagnosis System (RAMDS)

**Figure 2.**
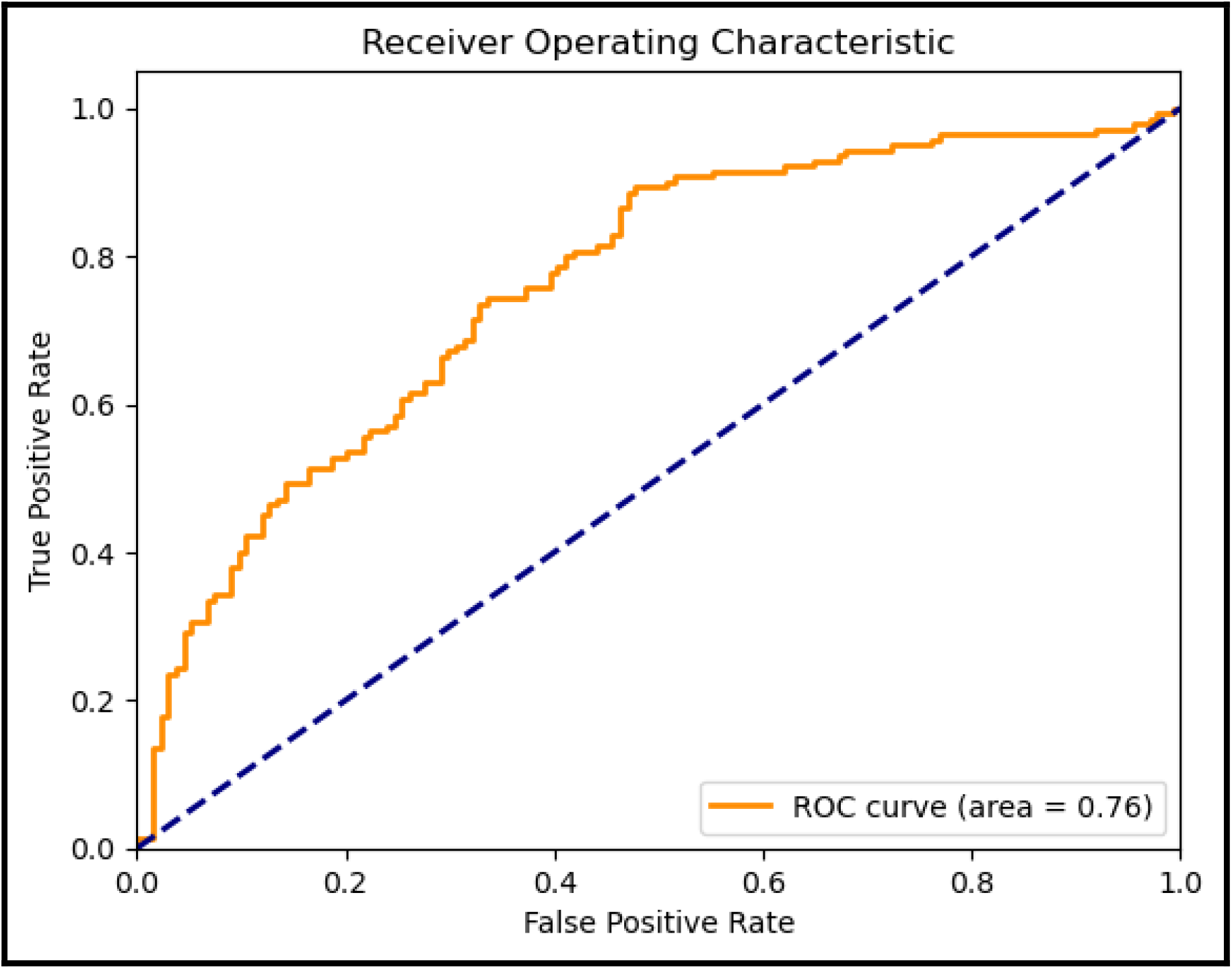
AUC ROC curve for the ResNet 34 model shows an area under the curve of 0.76.

### RAMDS results

Sensitivity, specificity, accuracy, positive predictive value, and negative predictive value for the retrieval augmented model were 90%, 49%, 70%, 65%, and 82% respectively. Since the retrieval augmented model only gave binary outputs, AUC couldn’t be calculated.

**Table 2.**
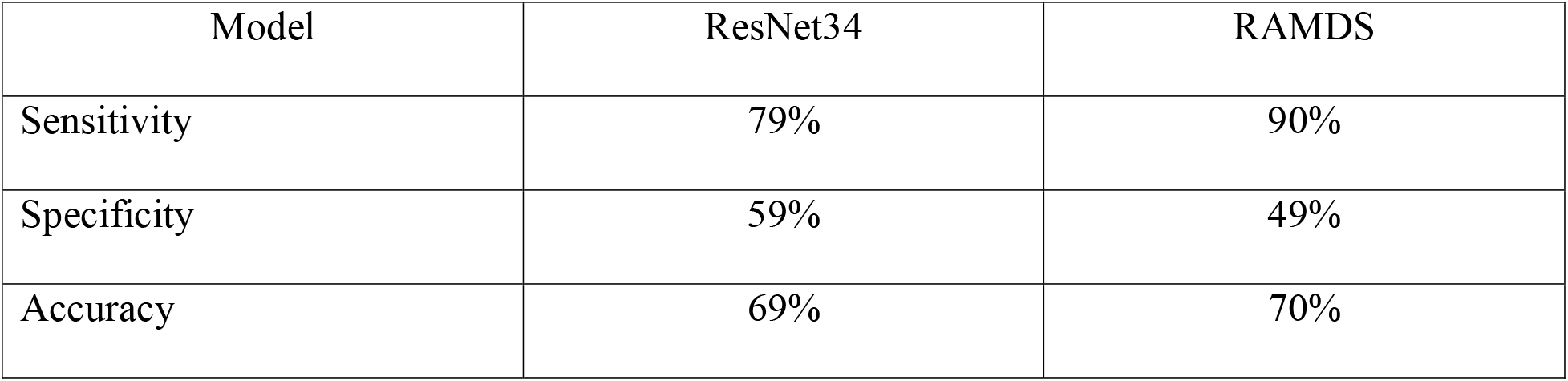
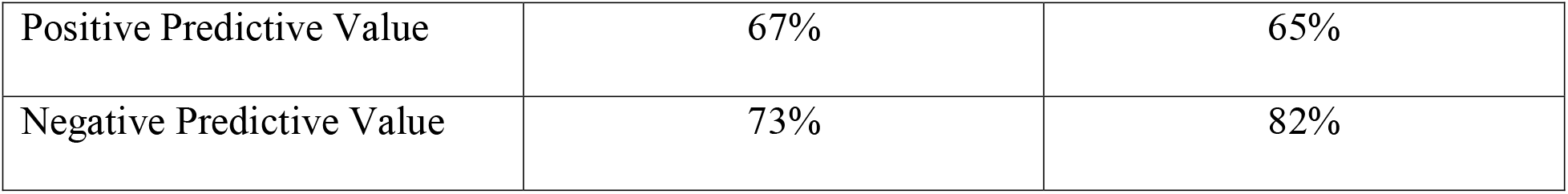
Comparing ResNet34 and RAMDS results.

| Model | ResNet34 | RAMDS |
| --- | --- | --- |
| Sensitivity | 79% | 90% |
| Specificity | 59% | 49% |
| Accuracy | 69% | 70% |
| Positive Predictive Value | 67% | 65% |
| Negative Predictive Value | 73% | 82% |

## Discussion

The Retrieval-Augmented Medical Diagnosis System (RAMDS) for breast cancer, employing machine learning and similarity-based algorithms, represents an advancement in the field of medical imaging and diagnosis. This system, integrating a pre-trained ResNet 34 model with a retrieval-augmented mechanism, demonstrates improved diagnostic performance while addressing the critical aspect of explainability in AI-driven medical decision-making when compared to the ResNet 34 model. The test data had a cancer prevalence of 51%. In real life screening, prevalence is much lower, which should theoretically enhance the negative predictive value of RAMDS system. RAMDS improved sensitivity by 11%, and negative predictive value by 9% when compared to the base model. This is comparable to having an experienced doctor who remembers every case that has been seen before and uses that knowledge to make better decisions.

### Explainability in AI Diagnostics

The challenge of explainability in AI-based medical diagnosis systems is a topic of considerable interest and concern in the medical community. Traditional deep learning models, while effective, often operate as “black boxes,” offering little insight into the reasoning behind their predictions (Castelvecchi, 2016). In contrast, the retrieval-augmented approach in this study enhances explainability by relating the AI’s diagnosis to similar historical cases. This approach aligns with the findings of Holzinger et al. (2017), who emphasized the importance of making AI decisions transparent, understandable, and explainable, especially in healthcare. This approach is also helpful in teaching less experienced operators using similar images.

The system’s reliance on similarity assessment with historical cases provides clinicians with a reference point, thereby facilitating a better understanding of the model’s decision-making process. This is very similar to the principles of case-based reasoning in medical diagnosis, as discussed by Bichindaritz and Marling (2006), where past cases are often used to inform the diagnosis of new cases.

Clinicians can also review similar images and corresponding diagnosis using RAMDS graphical user interface. After reviewing this information, they can decide whether to accept or reject the provided predictions based on how similar the images are. This makes clinicians an active participant and a partner in the AI assisted diagnostic model rather than a passive receiver of information.

### Adaptability to Local Imaging Contexts

An intriguing aspect of this system is its adaptability to different imaging contexts without the need for retraining the entire model. This is particularly relevant in medical imaging, where datasets can vary significantly in quality and characteristics across different institutions and regions (Shen et al., 2017). By creating embeddings for local images and fine-tuning the retrieval augmentation system, the model can be adapted to new contexts, without retraining the base model, enhancing its utility and scalability. This approach echoes the findings of Cheplygina et al. (2019), who highlighted the challenges of transferring AI models to new medical imaging datasets.

### Comparative Analysis with Existing Techniques

The performance metrics of the retrieval-augmented model, particularly in terms of sensitivity and negative predictive value, show a notable improvement over the base ResNet34 model. However, the specificity of the retrieval-augmented model was lower than that of the base model, which is an area that warrants further investigation. The trade-off between sensitivity and specificity in medical diagnosis models has been a subject of ongoing discussion, as highlighted by Lim (2021). When compared to previous literature on the positive predictive value of ultrasound in breast cancer diagnosis, RAMDS showed a large improvement. According to Berg and colleagues (Berg, 2015) positive predictive value (PPV) of breast ultrasound was 4.3%. RAMDS had a PPV of 65%.

### Limitations of the study

RAMDS was not tested in real world clinical settings. This hasn’t undergone regulatory scrutiny or approval. Prospective real-world testing is preferred when compared to retrospective testing on historical cases. The feasibility of incorporating RAMDS in the radiology workflow is critical and this has not been done. When making a decision to biopsy, radiologists review multiple images of the same lesion, but RAMDS makes predictions based on a single image. A future iteration of RAMDS could address these issues.

### Future Directions and Improvements

Future research could focus on enhancing the specificity of the retrieval-augmented model without compromising its sensitivity. Additionally, incorporating video clips, multi-modal data, such as combining ultrasound and mammographic findings could potentially improve diagnostic accuracy.

## Conclusion

Retrieval-augmented medical image diagnosis system presents a promising approach in the field of breast cancer diagnosis. Its strengths lie in its improved accuracy, adaptability, and particularly in its contribution to the explainability of AI-driven diagnoses. Further research and development are necessary to refine this system and fully realize its potential in clinical settings.

## Data Availability

BUSC_Mendeley - https://data.mendeley.com/datasets/wmy84gzngw/1
-Rodrigues, Paulo Sergio (2017), Breast Ultrasound Image, Mendeley Data, V1, doi: 10.17632/wmy84gzngw.1
BUSI_corrected - https://www.kaggle.com/datasets/jarintasnim090/busi-corrected
- Al-Dhabyani W, Gomaa M, Khaled H, Fahmy A. Dataset of breast ultrasound images. Data in Brief. 2020 Feb;28:104863. DOI: 10.1016/j.dib.2019.104863.
RODTOOk - origin - http://www.onlinemedicalimages.com/index.php/en/site-map
-Rodtook, A., Kirimasthong, K., Lohitvisate, W., Makhanov, S.S. (2018) Automatic initialization of active contours and level set method in ultrasound images of breast abnormalities. Pattern Recognition, Vol 79, pp 172-182".
QAMEBI - origin - https://qamebi.com/breast-ultrasound-images-database/
- [1] A. Abbasian Ardakani, A. Mohammadi, M. Mirza-Aghazadeh-Attari, U.R. Acharya, An open-access breast lesion ultrasound image database‏: Applicable in artificial intelligence studies, Computers in Biology and Medicine, 152 (2023) 106438. https://doi.org/10.1016/j.compbiomed.2022.106438
- [2] H. Hamyoon, W. Yee Chan, A. Mohammadi, T. Yusuf Kuzan, M. Mirza-Aghazadeh-Attari, W.L. Leong, K. Murzoglu Altintoprak, A. Vijayananthan, K. Rahmat, N. Ab Mumin, S. Sam Leong, S. Ejtehadifar, F. Faeghi, J. Abolghasemi, E.J. Ciaccio, U. Rajendra Acharya, A. Abbasian Ardakani, Artificial intelligence, BI-RADS evaluation and morphometry: A novel combination to diagnose breast cancer using ultrasonography, results from multi-center cohorts, European Journal of Radiology, 157 (2022) 110591. https://doi.org/10.1016/j.ejrad.2022.110591

## Appendix

### Appendix I: RAMDS user interface

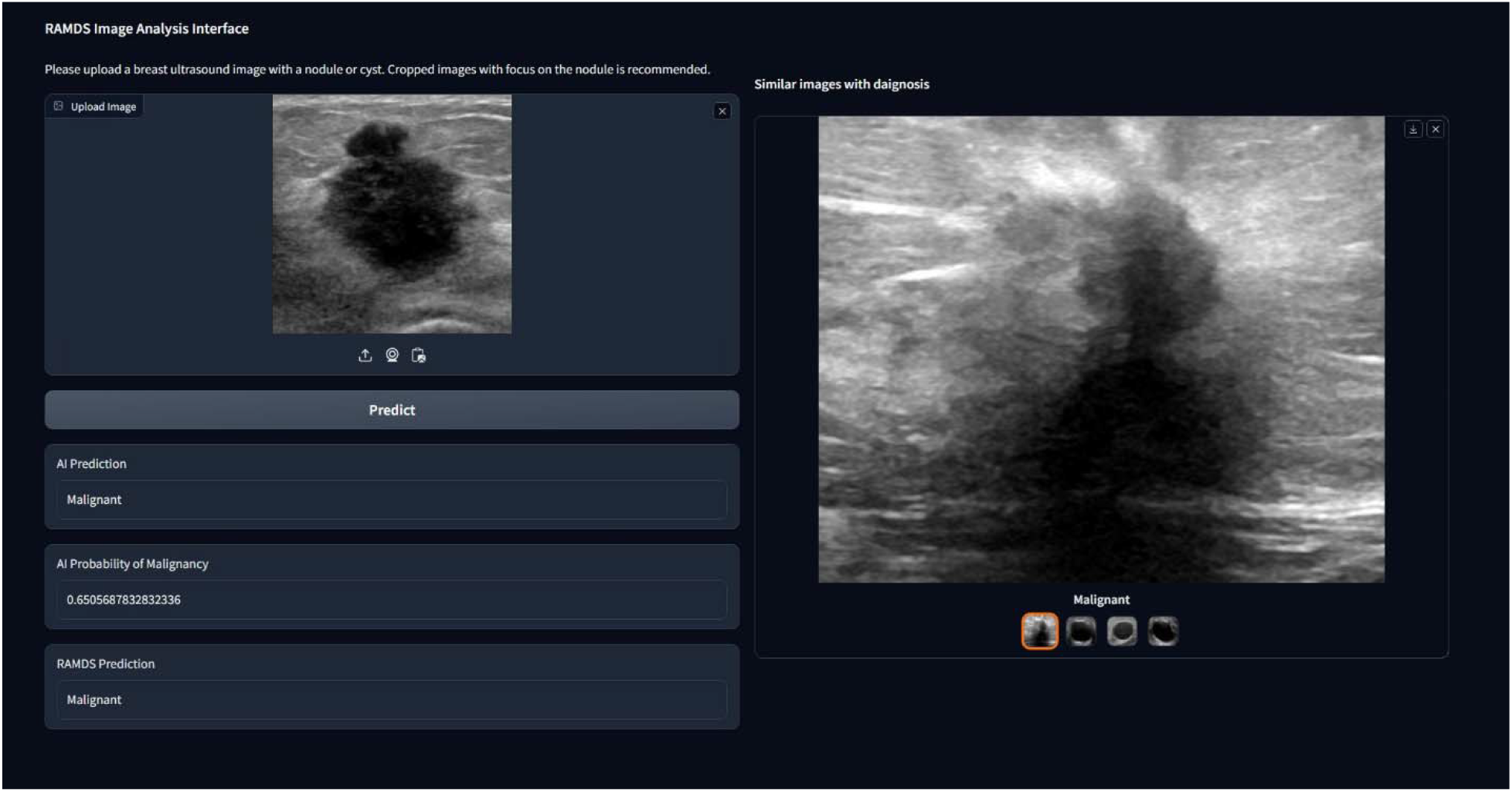

User interface for RAMDS.

Users can upload an ultrasound image as seen on the left side of the picture. Once the image is uploaded, users can click the predict button to populate the AI Prediction, AI Probability of Malignancy, RAMDS Prediction text fields, and the similar images picture gallery. Similar images picture gallery shows images that are similar to the uploaded image from the training dataset along with the corresponding diagnosis.

